# Deep learning-derived age of hippocampus-centred regions is influenced by *APOE* genotype and modifiable risk factors

**DOI:** 10.1101/2024.10.27.24316212

**Authors:** Chao Dong, Yuangang Pan, Anbupalam Thalamuthu, Jiyang Jiang, Jing Du, Karen A. Mather, Perminder S. Sachdev, Wei Wen

**Affiliations:** Centre for Healthy Brain Ageing (CHeBA), Discipline of Psychiatry and Mental Health, School of Clinical Medicine, UNSW, Sydney, Australia; Centre for Frontier AI Research (CFAR), A*STAR, Singapore; Neuropsychiatric Institute (NPI), Prince of Wales Hospital, Randwick, New South Wales, 2031, Australia; Department of Psychology, University of Cambridge, Cambridge, CB2 3EB, United Kingdom

**Keywords:** hippocampus, brain age, deep learning, convolutional neural network, *APOE*

## Abstract

Brain age has been widely investigated by using the whole brain image. However, the age of some specific brain regions, such as those related to the hippocampus, remains underexplored. This study developed age prediction models for left and right hippocampus-centred regions of interest (hippocampus ROI) using three-dimensional convolutional neural networks (3D-CNN) based on MRI scans from 31,370 healthy participants in the UK Biobank. The hippocampus ROI age (HA) gap was calculated by subtracting chronological age from predicted HA. Additionally, the longitudinal change rate of the HA gap was estimated in 3,893 participants with imaging data at two time points over an average follow-up of 2.63 years. The models achieved state-of-the-art performance (mean absolute error (MAE): 2.47 – 2.84 years). Cross-sectional analysis revealed that *APOE* ε4 homozygotes had a greater HA gap compared to *APOE* ε4 non-carriers. Participants with hypertension, diabetes, heavy alcohol consumption, or smoking also exhibited larger HA gap. Transfer learning applied to an independent dataset confirmed similar trends in some variables, though findings were not statistically significant. Interestingly, longitudinal analysis showed that *APOE* ε4 homozygotes had a higher annual change rate in the left HA gap compared to *APOE* ε2 homozygotes. Occlusion analysis saliency maps indicated that regions around the hippocampus, including the thalamus, pallidum, nearby cerebral cortex, and white matter, significantly contributed to the age prediction. The left HA gap emerges as a potential biomarker linked to the *APOE* genotype and an indicator of health.

## 1 Introduction

Medial temporal lobe (MTL), which includes the hippocampus, entorhinal, perirhinal, and parahippocampal cortices, is altered early and severely in Alzheimer’s disease (AD) (Chauveau et al., 2021; de Flores et al., 2020) and is also one of risk factors for vascular dementia (Pendlebury and Rothwell, 2009). AD is characterised by initial regional brain atrophy in the hippocampus and medial temporal regions, followed by subsequent spread to other cortical areas (Chapleau et al., 2016). The temporal lobes and hippocampus are high-risk regions for AD classification (Qiu et al., 2020; Wang et al., 2023), or related dementia (Martin et al., 2023). Therefore, an in-depth exploration of hippocampus-centred brain regions could provide great neuroscientific value.

The Apolipoprotein E (*APOE*) gene, which is polymorphic with three common isoforms: ε2, ε3, and ε4, is encoded by two single nucleotide polymorphisms. *APOE* ε4 is the strongest genetic risk factor for late-onset AD, *APOE* ε3 is neutral, and *APOE* ε2 is protective (Huang et al., 2017; Serrano-Pozo et al., 2021). Individuals with one copy of *APOE* ε4 (*APOE* ε4 heterozygotes) have an increased risk of developing AD, and those with two copies ( *APOE* ε4 homozygous) have an even higher risk (Gharbi-Meliani et al., 2021). *APOE* ε4 has been associated with brain atrophy (Cacciaglia et al., 2018), white matter lesions, memory decline (Koizumi et al., 2018), and MTL subregions (de Flores et al., 2023). In addition to AD, *APOE* ε4 has also been associated with vascular dementia (Skrobot et al., 2016), Parkinson’s disease and Lewy body dementia (Rongve et al., 2019; Saeed et al., 2021).

In longitudinal studies involving patients with AD, the rate of hippocampal atrophy is influenced by both *APOE* genotypes and the severity of the disease, with *APOE* ε4 carriers showing greater rates of atrophy (Mori et al., 2002). *APOE* ε4/ε4 homozygotes also exhibited accelerated atrophy in hippocampus (Abushakra et al., 2020). Previous study suggests that *APOE* ε4 carriers may exhibit a greater rate of hippocampal atrophy compared to their counterparts, even without an AD diagnosis (Moffat et al., 2000). Furthermore, *APOE* ε4 carriers also showed faster rates of structural loss in MTL (Donix et al., 2010; Mishra et al., 2018). Additionally, accumulating evidence showed that hypertension, diabetes, excessive alcohol consumption, smoking, and physical inactivity are high risk factors for dementia or adverse brain health outcomes (Chudasama et al., 2020; Franz et al., 2021; Gottesman and Seshadri, 2022; Livingston et al., 2020). These modifiable risk factors can influence hippocampal atrophy (Fotuhi et al., 2012) and multiple body age (Tian et al., 2023).

Brain age, or brain-predicted age, can be estimated using neuroimaging data and may differ from chronological age, resulting in a brain age gap (Cole and Franke, 2017). By summarizing complex neuroimaging features into a simple, interpretable summary metric, brain age gap may reflect a comprehensive indicator of personalized brain health, providing prognostic and predictive values (Cole and Franke, 2017; Millar et al., 2023). For example, a greater brain age gap is associated with AD (Gaser et al., 2013) and mortality (Cole et al., 2018). Grey matter density around hippocampus and amygdala has been identified as a key factor influencing age predictions (Wang et al., 2019). A recent study which used the hippocampal-centred regions to predict brain age found that brain age gap might be a biomarker to help AD and mild cognitive impairment (MCI) diagnosis (Poloni and Ferrari, 2022). Cumulatively, this evidence highlights the critical importance of the hippocampus and its neighbouring structures. Additionally, a study demonstrated that the 3D bounding boxes of the ROI surrounding the hippocampus could act as a promising representation for dementia diagnosis and prediction in deep learning models, as it contains information around hippocampus structure and there is no requirement of the accurate hippocampus segmentation (Sarasua et al., 2022). However, most studies on brain age are cross-sectional, and longitudinal studies are needed to explore the change rate of the predicted age.

In this study, we conceptualized the age of “hippocampus-centred regions of interest (hippocampus ROI age)” as a measure representing region-specific biological age. Hippocampus ROI age (HA) can be regarded as the proxy for the biological age of the hippocampus ROI which are 3D bounding boxes centred around hippocampus. They do not only contain structure information of hippocampus but also their nearby brain structures. Here, (1) we aimed to estimate the deep learning-based HA from 31,370 healthy participants who aged 44 to 83 (five-fold cross-validation) and explore whether the presence of the *APOE* ε4 allele and modifiable risk factors are associated with HA gap. (2) We aimed to evaluate the longitudinal change rate of the HA gap in different *APOE* genotype groups. (3) We also aimed to identify which areas contribute more to the HA prediction by applying occlusion analysis. As the hippocampus ROI are the first to be affected by AD, and *APOE* ε4 is the major genetic risk factor for AD, we hypothesised that HA gap might be associated with *APOE* genotype, with *APOE* ε4/ ε4 carriers exhibiting a greater HA gap.

## 2 Methods

### 2.1 Participants

*UK Biobank*. The UK Biobank is a large-scale dataset containing cross-modality neuroimaging data. It is a population-based study which consists of over 500,000 participants aged between 40-70 years at study entry (Sudlow et al., 2015). Written consent was acquired from all participants and ethics approval was provided by the National Health Service National Research Ethics Service (11/NW/0382). In the current study, after excluding participants with severe self-reported brain related disorders (Table S1), 31,370 healthy participants with one time point image were included in the final analysis. Additionally, there were 3,893 participants who have two-time points image. All the 31,370 healthy participants with one time point image were randomly split into five equally sized folds, with 60% for training, 20% for validation, and 20% for test set. Model performance in the training set was assessed using 5-fold cross validation.

*Sydney Memory and Ageing Study (MAS)*. MAS is a cohort of 1,037 community dwelling adults aged 70–90 years (Sachdev et al., 2010). Ethics approval was obtained from the Human Research Ethics Committees of the University of New South Wales and the South Eastern Sydney Local Health District. For the current study, 534 participants with MRI scans from baseline were used.

### 2.2 Non-imaging data

#### 2.2.1 *APOE* genotype

For UK Biobank, DNA extracted from the blood samples of the initial cohort (∼ 50,000 participants), underwent genotyping within the UK Biobank utilising the Affymetrix UK BiLEVE Axiom array. For MAS, DNA was extracted from peripheral blood leukocytes or saliva samples using standard procedures at Genetics Repositories Australia (www.powmri.edu.au/GRA.htm) (Sachdev et al., 2010). Two *APOE* coding single nucleotide polymorphisms (SNPs), namely rs7412 and rs429358, were retrieved from the genotyped data to determine the *APOE* genotype. There are six *APOE* genotypes: ε2/ε2, ε2/ε3, ε2/ε4, ε3/ε3, ε3/ε4, and ε4/ε4. *APOE* ε4 allele carriers include ε2/ε4, ε3/ε4, and ε4/ε4. Non-carriers include ε2/ε2, ε2/ε3, and ε3/ε3.

#### 2.2.2 Cognitive measures

UKB: Cognitive assessments were administered on a fully automated touchscreen questionnaire (Fawns-Ritchie and Deary, 2020; Sudlow et al., 2015). Seven tests from the UK Biobank battery of cognitive tests were selected for the current study to represent three cognitive domains: “Reaction Time”, “Trail Making Test A”, and “Symbol Digit Substitution” formed the Processing Speed domain; “Numeric Memory” and “Pairs Matching” contributed to the Memory domain; and “Trail Making Test B” and “Fluid Intelligence” formed the Executive Function domain. All test scores were first z-transformed and then averaged to form domain scores. Therefore, processing speed, executive function, and memory were used in the current study. Global cognition was estimated by averaging the domain scores and z-transform.

MAS: A battery of interview-based cognitive tests was administered by trained psychologists to examine cognitive domains (Sachdev et al., 2010). Cognition domains include: attention processing speed (including “Digit Symbol-Coding” and “Trail Making Test A”), memory (including “Logical Memory Story A delayed recall”, “Rey Auditory Verbal Learning Test”, and “Benton Visual Retention Test recognition”), verbal memory (including “Logical Memory Story A delayed recall” and “Rey Auditory Verbal Learning Test”), language (including “Boston Naming Test – 30 items” and “Semantic Fluency (Animals)”), visuospatial (“Block Design”), executive function (“Controlled Oral Word Association Test (FAS)” and “Trail Making Test B”). All the details can be found in previous studies (Sachdev et al., 2010).

#### 2.2.3 Modifiable risk factors

In the current study, we focused on the following modifiable risk factors from UK Biobank: (1) alcohol intake frequency [Data-Field 1558], (2) smoking status [Data-Field 20116], (3) physical activity [Data-Field 22040], (4) hypertension [Data-Field 4079 and Data-Field 4080], and (5) diabetes [Data-Field 2443]. Specially, alcohol intake frequency was assessed based on questionnaire, and it was classified as: daily or almost daily, three or four times a week, once or twice a week, one to three times a month, special occasions only, or never. Smoking status was assessed by a questionnaire. Individuals were asked whether they were never, previous, or current smokers. Physical activity was measures by metabolic equivalent of task (MET) calculation (Doherty et al., 2017; Le Goallec et al., 2023). Data was self-reported and classified as: no physical activity, medium (>0, <7.5 MET hours/week), and medium (>0, <7.5 MET hours/week). Participants with hypertension (high blood pressure) were defined with blood pressure over 140/90 mmHg (https://www.who.int/news-room/fact-sheets/detail/hypertension). Diabetes was defined according to the doctor’s diagnosis. Additionally, we also considered body mass index (BMI) (kg/m^2^), which was calculated based on the measured weight and height: normal (18.5-24.9 kg/m^2^), underweight (<18.5 kg/m^2^), overweight (25.0-29.9 kg/m^2^), obese (> 30 kg/m^2^). In MAS, smoking status, hypertension, and diabetes are defined in the same way as in UK Biobank. Alcohol consumption was classified as never, previous, or current drinkers.

### 2.3 Image data and processing

UK Biobank structural MRI scans were acquired on three 3T Siemens Skyra MRI scanners. The key parameters for MRI imaging were (a) T1-weighted MRI: TI = 880 ms, TR = 2 000 ms, resolution = 1.0 × 1.0 × 1.0 mm, matrix size = 208 × 256 × 256; (b) T2-weighted FLAIR: TI = 1 800 ms, TR = 5 000 ms, resolution = 1.05 × 1.0 × 1.0 mm, matrix size = 192 × 256 × 256.The full protocol is provided at http://biobank.ctsu.ox.ac.uk/crystal/refer.cgi?id=2367. FreeSurfer (version 7.1.0) was used to model the cortical surface. For MAS cohort, we used T1-weighted and FLAIR scans and the key parameters for MRI imaging were: (a) T1- weighted MRI: TR = 6.39 ms, TE = 2.9 ms, flip angle = 8°, matrix size = 256×256, FOV (field of view) = 256×256×190, and slice thickness = 1 mm with no gap in between, yielding 1×1×1 mm^3^ isotropic voxels; (b) T2-weighted FLAIR: TR = 10000 ms, TE =110 ms, TI = 2800 ms, matrix size = 512×512, slice thickness = 3.5 mm without gap, and in plane resolution = 0.488×0.488 mm.

We used both the T1 and T2 FLAIR images as inputs to the FreeSurfer modelling (or only the T1 when the T2 was not available). The FreeSurfer output image “nu.mgz” was used for the extraction of hippocampus-centred regions in the current study, with a resolution of 1 × 1 × 1 mm and inhomogeneity correction applied. The process for defining hippocampus-centred regions involved the following steps:

1. The gravity centre of each participant’s left and right hippocampus was identified, and then the lower, upper, left, right, front, and back dimensions were extended by 32 voxels based on the gravity centre, resulting in hippocampus-cantered regions of interest (ROIs) measuring 64 × 64 × 64 voxels.
2. The reference brain in a standard space underwent the same method to obtain the standard space hippocampus ROI.
3. The hippocampus ROI in standard space served as a reference for registering each participant’s hippocampus ROI, obtaining the registration transformation matrix.
4. The transformation matrix was applied to register the entire brain image, followed by repeating step (1) to acquire the final hippocampus ROI with a size of 64 × 64 × 64 voxels.

By following these procedures, we achieved the consistency of each participant’s left and right hippocampus ROI in the same space.

### 2.4 3D-CNN architecture

The architecture follows the Simple Fully Convolutional Network (SFCN) architectures (Peng et al., 2021). The architecture consisted of six 3D convolutional layers with varying channel numbers [32, 64, 128, 256, 256, 64]. Each convolutional layer has a kernel size of 3, padding of 1, and a stride of 2. Following the convolutional layers is a two-layer Multilayer Perceptron (MLP). The input data size was 64 × 64 × 64, and the output was the predicted hippocampus ROI age in years. Specifically, each of the first five 3D convolutional layer was followed by a batch normalisation layer, a max pooling layer, and a Rectified Linear Unit (ReLU) activation layer. The sixth 3D convolutional layer was followed solely by a ReLU activation layer, without a max pooling layer. The dimension of the output after all convolutional layers was 512. Subsequently, the output from the convolutional layers was flattened to be fed into two MLP layers. The first fully connected (linear) layer produced 100 output features, and the second produced a single scalar value, representing the final predicted age. During training, a learning rate of 1×10^−3^ was employed, and the batch size was set to 128.

Predicted ages often face challenges such as underfitting caused by regression dilution and a non-Gaussian age distribution. This implies that older participants might be estimated with a younger brain age, while younger participants might be estimated with an older brain age. Bias correction is an essential postprocessing technique in most brain-age prediction studies, and age correction was performed using method described in Smith et al. study (Smith et al., 2019). We fitted a linear regression y2=a×y1+b (y1: chronological age, y2: predicted age) to the validation set, and then applied the learned coefficients (a, b) to the test set. Therefore, the corrected brain age can be calculated by (y2-b)/a. All further analyses used the age-corrected gap. To avoid inflating estimates of prediction accuracy (Butler et al., 2021), the uncorrected predicted age values were applied for model performance evaluation. Model performance was evaluated by mean absolute error (MAE) and person’s correlation between chronological age and predicted age.

Additionally, we also applied the whole brain image (FreeSurfer output image “nu.mgz”) to the 3D-CNN architecture to predict the whole brain age. Details can be found in supplementary materials.

### 2.5 Statistical analysis

#### Cross-sectional analysis

All statistical analyses were conducted in R 4.0.2. Participants’ predicted age was merged across the 5 folds, so that each participant can get a predicted hippocampus-centred regions age.

##### The HA gap in *APOE* genotype groups

We investigated the HA gap estimation in the participants with *APOE ε2/ε2, APOE ε2/ε3, APOE ε3/ε3, APOE ε3/ε4,* and *APOE ε4/ε4* genotype. We also grouped participants as *APOE ε4* homozygotes (*ε4/ε4*), *APOE ε4* heterozygotes (*ε2/ε4, ε3/ε4*), and *APOE ε4* non-carriers. HA gap was set as the dependent variable, and *APOE ε4* carrier status (or *APOE* genotype), age, sex, scanner, and intracranial volume (ICV) were set as independent variables. Similarly, we also investigated the associations between hippocampus volume and *APOE ε4* carrier status (or *APOE* genotype) using similar regression model by setting hippocampus volume (HV) as the dependent variable. For both analyses, *APOE* ε4 non-carriers served as the reference group, with other groups compared against this reference. Additionally, *APOE* genotypes ε2/ε2, ε2/ε3, and ε3/ε3 were individually set as reference groups for comparisons with the remaining groups.

##### Associations between HA gap and cognition

We also performed regression model to explore the associations between HA gap and cognition, by setting cognition as the dependent variable and HA gap as independent variable. All variables were z-transformed. We also included chronological age, sex, scanner, and years of education. as covariance variables in the regression model. Additionally, associations between hippocampus volume and cognition were calculated using a similar method.

##### HA and modifiable risk factors

Associations between the left HA gap and modifiable risk factors (hypertension and diabetes, alcohol intake frequency, smoking status, physical activity) were explored by applying the regression model: left or right HA gap ∼ hypertension + diabetes + alcohol intake frequency + smoking status + physical activity + BMI + *APOE* ε4 status + baseline age + sex + scanner + ICV. Here, BMI, *APOE* ε4 status, baseline age, sex, scanner, and ICV were used as covariance variables. Each risk factor had a specific reference group: participants who are non-hypertensive, or non-diabetic, drink daily or almost daily, those who have never smoked, engage in no physical activity. Other corresponding groups were compared against these reference groups individually.

#### Longitudinal analysis

##### The HA gap change rate in *APOE* genotype groups

The annual HA gap change rate was calculated by the gap difference between baseline and follow-up and then divided by the time interval between these two time points (years). The change rate was compared within different *APOE* genotype groups. We applied the similar regression model, which was used in the cross-sectional analysis, but set annual HA gap change as the dependent variable instead of HA gap. The annual hippocampus volume change was also estimated in a similar regression model. All variables were z-transformed, and these regression models were controlled by baseline age, sex, scanner, and ICV.

### 2.6 Occlusion analysis

To interpret the CNN model results and understand the regions of the input that contribute HA prediction, occlusion analysis (Lee et al., 2022; Zeiler and Fergus, 2014) was conducted within the test dataset. Specifically, the input images were divided by 4×4×4 grid, and for each square region, a corresponding occlusion mask with 16×16×16 voxels was applied. The occlusion process involved systematically making the different occlusion mask with zero values. The age prediction on occluded images was performed using our original pretrained CNN model, and the model performance was evaluated by MAE_occlusion_. The delta MAE was calculated as MAE_delta_ = MAE_occlusion_ - MAE_original_ (MAE_original_ represents the MAE of CNN model using original images). A delta MAE matrix (4×4×4) was generated by iterating occlusion for each square region. Finally, cubic interpolation was applied to the delta MAE matrix to obtain the original image size (64×64×64). This procedure was done in each test set in five-fold and the average of the 5 folds was finally calculated. Normalization was carried out by dividing the whole image by its maximum value.

To examine how important the hippocampus is in the HA prediction, we also removed hippocampus from the original ROI (occluded using the hippocampus mask), and then used it as the input of the pretrained model. The MAE was estimated to assess the model’s performance.

### 2.7 Apply the 3D-CNN on a new dataset using transfer learning

We continue to examine the 3D-CNN model performance on another dataset: MAS cohort. As the sample size in MAS dataset is relatively small (N=523), we used 3-fold cross validation within MAS dataset, with 2/3 participants using as the training set and 1/3 as test set. The imaging processing procedure was the same as the procedure in UKB dataset. Specifically, the MAS dataset was randomly split into 3 folds. The MAS training set was full-fined using the pre-trained 3D-CNN model in UKB. As we used 5-fold cross validation in UKB, these 5 pre-trained models were applied to transfer learning separately.

## 3 Results

### 3.1 Demographics

Table 1 summarizes the demographic characteristics of the participants from the UK Biobank and the MAS. In the current study, 31,370 participants in UK Biobank were included in the five-fold analysis, and 26,206 of them have both HA data and genetic data, with *APOE* ε4 non-carriers (72.18%), *APOE* ε4 heterozygotes (25.59%), and *APOE* ε4 homozygotes (2.23%). In the longitudinal analysis, 3,276 participants were included, with *APOE* ε4 non-carriers (72.92%), *APOE* ε4 heterozygotes (24.97%), and *APOE* ε4 homozygotes (2.11%). The MAS cohort included 523 participants with a mean age of 78.35 years (SD = 4.66). The study workflow can be found in Figure 1.

**Figure 1.**
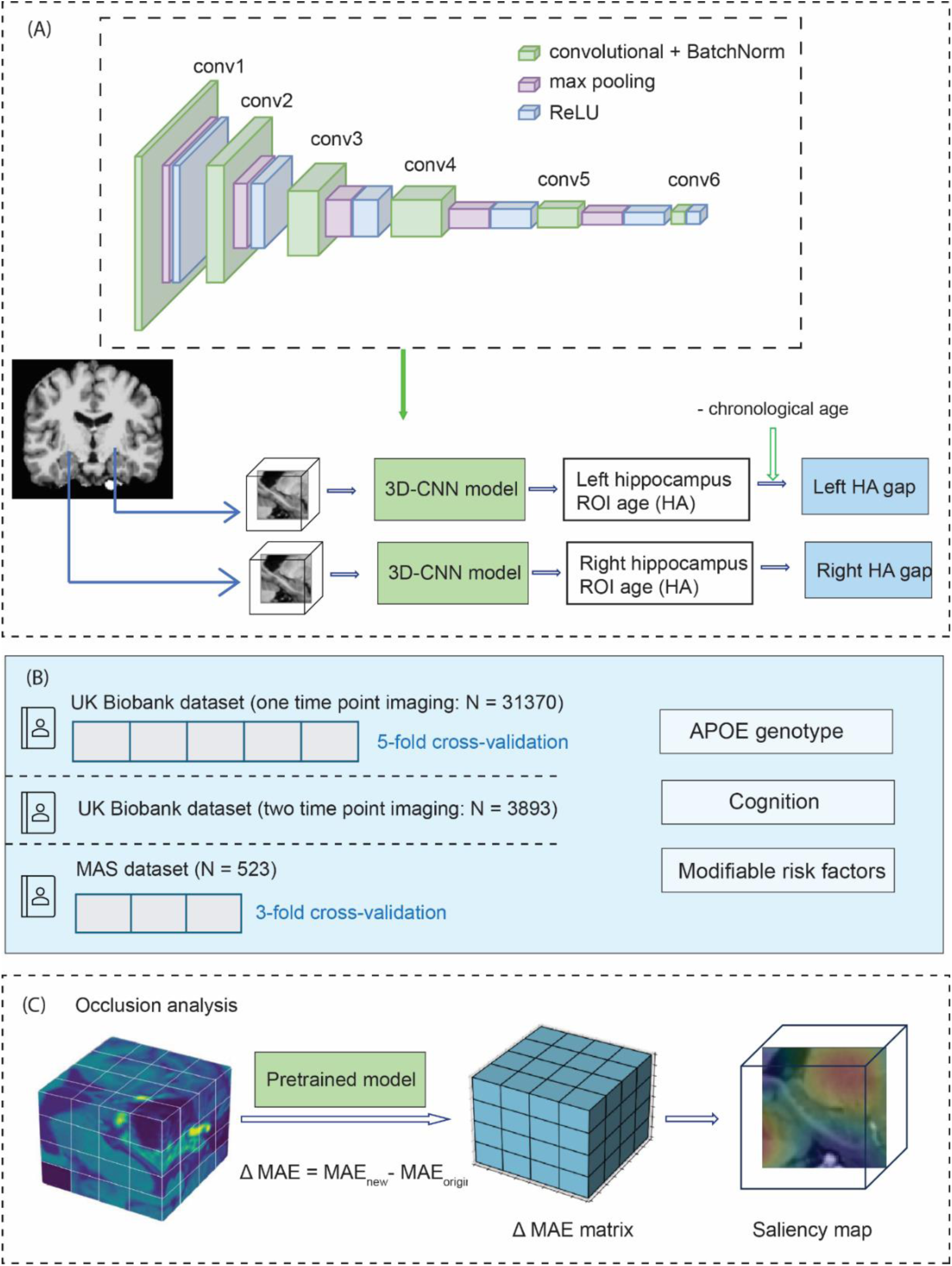
Study workflow. The architecture of the age prediction model in left and right hippocampus-centred regions of interest (hippocampus ROI) based on three-dimensional convolutional neural network (3D-CNN). Model input was 3D 64 × 64 × 64 voxels hippocampus ROI image, and output was the predicted hippocampus ROI age (HA). The HA gap was calculated by subtracting the chronological age from the HA. Associations between HA gap and APOE genotype, cognition, and modifiable risk factors were further explored. The pre-trained model from the UK Biobank dataset was applied to the MAS dataset using transfer learning. (B) Dataset in the current study. (C) Occlusion analysis. MAS: Sydney Memory and Ageing Study.

**Table 1.** Demographic characteristics.

|  |  |  |  |
| --- | --- | --- | --- |
| Participants<br>(number) | **UKB:**<br>Only have one time<br>point imaging data.<br>(N=31,370) | **UKB:**<br>Have two-time points<br>imaging data.<br>(N=3,893) | **MAS:**<br>(N=523) |
| Age, years, mean<br>(SD) | 64.50 (7.52) | Baseline: 62.07 (7.40)<br>Follow-up: 64.70<br>(7.20)<br>Gap years: 2.63 (1.05) | 78.35 (4.66) |
| Male, number (%) | 14915 (47.55%) | 1816 (46.65%) | 233 (44.55%) |
| **Genetic Data** |  |  |  |
| **APOE genotype,**<br>number (%) | Total: N=26,206 | Total: N=3,276 | Total: N=517 |
| $\epsilon 2\epsilon 2$ | 135 (0.52%) | 19 (0.5%) | 3 (0.58%) |
| $\epsilon 2\epsilon 3$ | 3235 (12.34%) | 393 (12%) | 74 (14.31%) |
| $\epsilon 2\epsilon 4$ | 605 (2.31%) | 88 (2.69%) | 6 (1.16%) |
| $\epsilon 3\epsilon 3$ | 15546 (59.32%) | 1977 (60.35%) | 316 (61.12%) |
| $\epsilon 3\epsilon 4$ | 6101 (23.28%) | 730 (22.28%) | 109 (21.08%) |
| $\epsilon 4\epsilon 4$ | 584 (2.23%) | 69 (2.11%) | 9 (1.74%) |
| $\epsilon 4$ non-carriers | 18916 (72.18%) | 2389 (72.92%) | 393 (76.02%) |
| $\epsilon 4$-heterozygotes | 6706 (25.59%) | 818 (24.97%) | 115 (22.24%) |
| $\epsilon 4$-homozygotes | 584 (2.23%) | 69 (2.11%) | 9 (1.74) |

**Notes:** UKB: UK Biobank. MAS: Sydney Memory and Ageing Study. SD: standard deviation.

### 3.2 Cross-sectional analysis

#### The HA gap in *APOE* genotype groups

Our model achieved a competitive state-of-the-art metric (MAE: 2.52 – 2.84 years in left HA model; 2.47 – 2.72 years in right HA model) (Table 2), with the correlation between chronological and predicted age ranging from 0.901 to 0.907 (Figure S1). Compared with *APOE* ε4 non-carriers, *APOE* ε4 homozygotes (ε4/ε4) showed significantly greater HA gap (left: β = 0.166, 95% confidence interval [CI] = [0.085, 0.248], p = 6.30e-05; right: β = 0.192, CI = [0.111, 0.274], p = 4.03e-06) and smaller hippocampus volume in both left and right hemispheres. Specifically, the HA gap in *APOE* ε4 homozygotes group was also significantly greater than that in ε2/ε2, ε2/ε3 and ε3/ε3 group (Figure 2 and Table 3).

**Figure 2.**
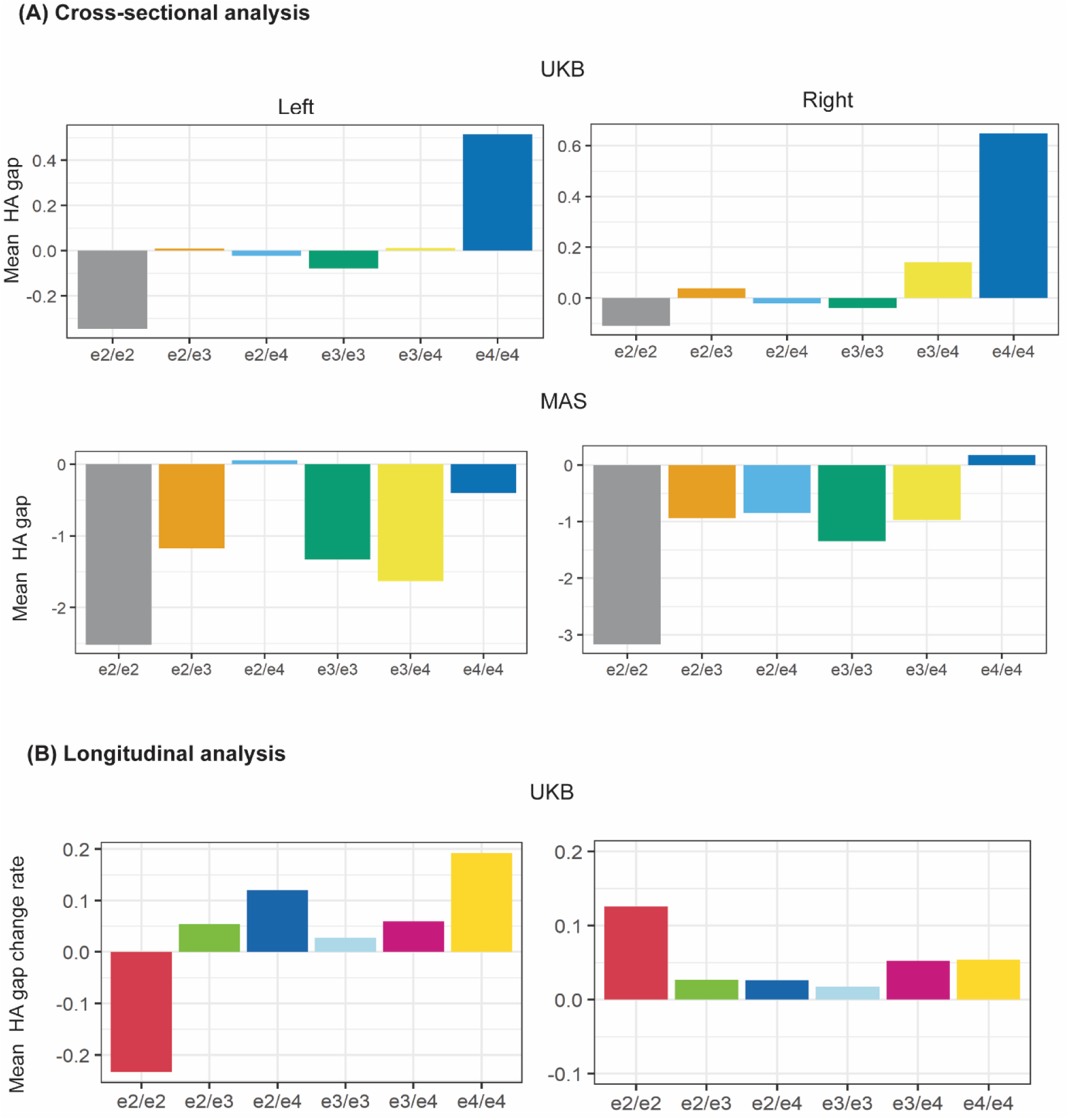
Left and right mean HA gap in cross-sectional analysis and mean HA gap change rate in longitudinal analysis based on *APOE* genotype. (A) Cross-sectional analysis in UKB and MAS. (B) Longitudinal analysis in UKB. X axis represents six *APOE* genotypes, and Y axis represents mean HA gap value (cross-sectional) or mean HA gap change rate (longitudinal). UKB: UK Biobank. MAS: Sydney Memory and Ageing Study. HA: hippocampus ROI age.

**Table 2.**
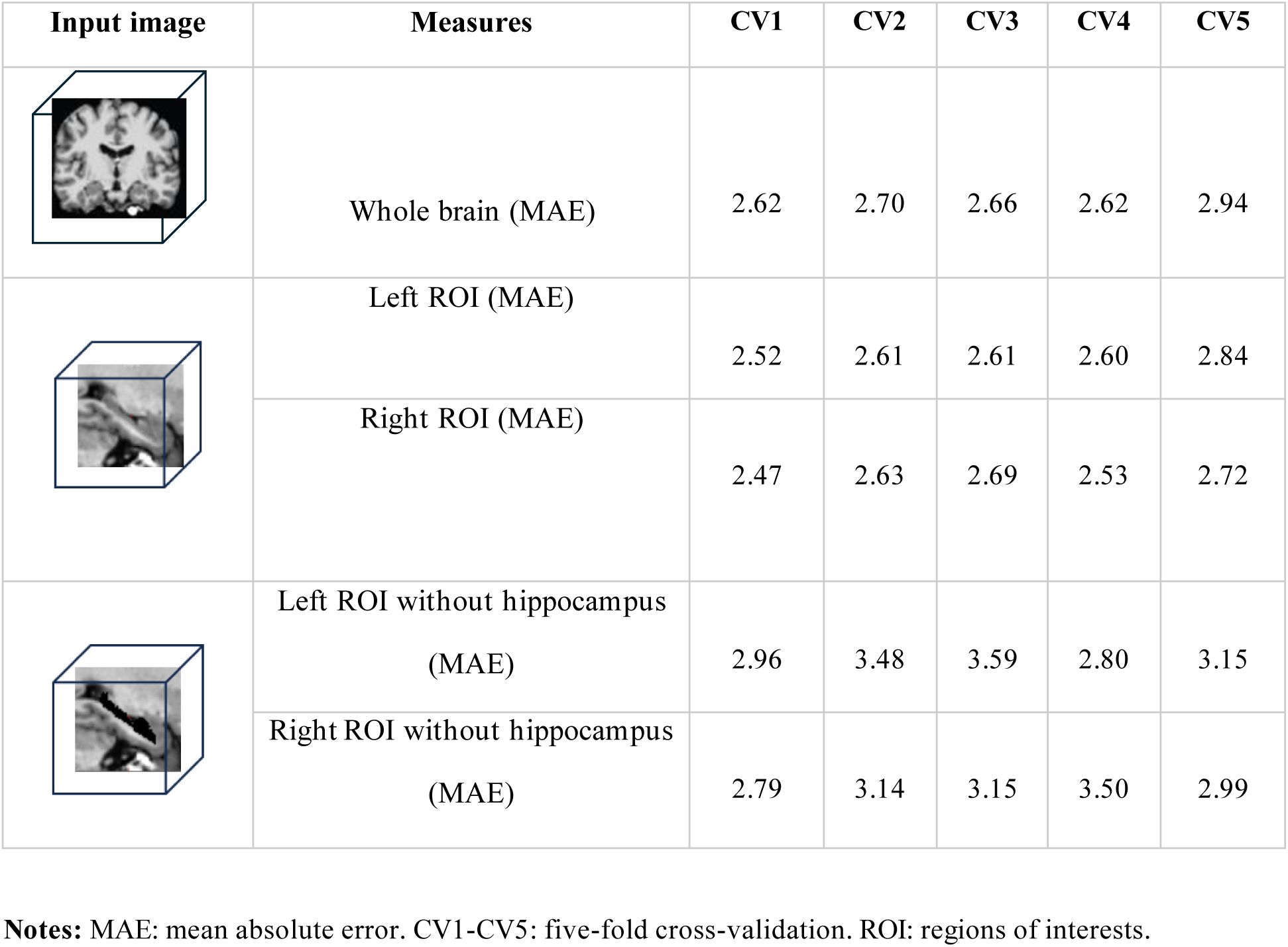
Performance of 3D Convolutional Neural Network (3D-CNN) model with five-fold cross validation (evaluated by MAE) in the UK Biobank.

| Input image | Measures | CV1 | CV2 | CV3 | CV4 | CV5 |
| --- | --- | --- | --- | --- | --- | --- |
|  | Whole brain (MAE) | 2.62 | 2.70 | 2.66 | 2.62 | 2.94 |
|  | Left ROI (MAE) | 2.52 | 2.61 | 2.61 | 2.60 | 2.84 |
|  | Right ROI (MAE) | 2.47 | 2.63 | 2.69 | 2.53 | 2.72 |
|  | Left ROI without hippocampus (MAE) | 2.96 | 3.48 | 3.59 | 2.80 | 3.15 |
|  | Right ROI without hippocampus (MAE) | 2.79 | 3.14 | 3.15 | 3.50 | 2.99 |

**Notes:** MAE: mean absolute error. CV1-CV5: five-fold cross-validation. ROI: regions of interests.

**Table 3:**
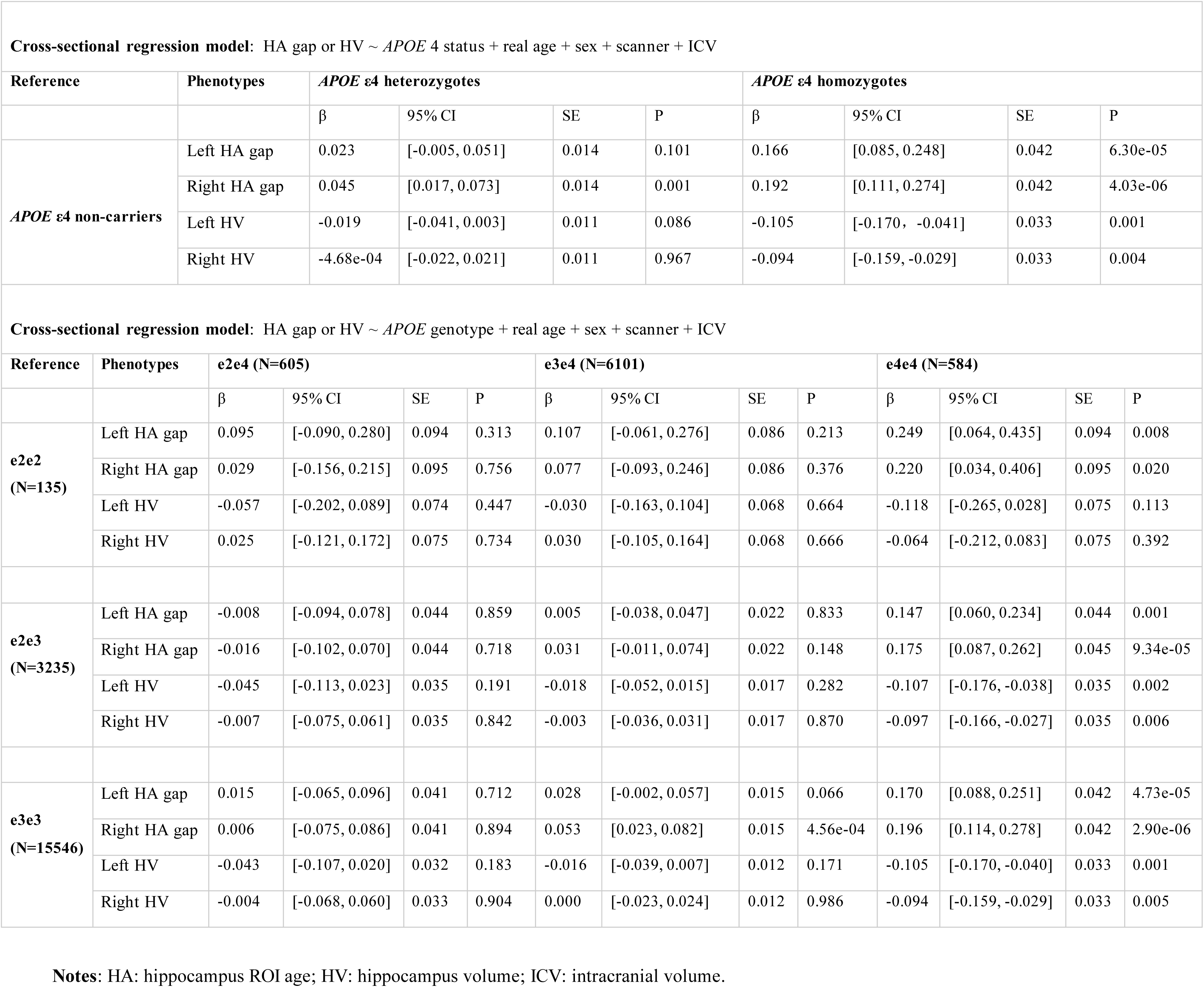
Cross-sectional analysis in UK Biobank: The HA gap and HV in different *APOE* genotype groups.

#### Whole brain age gap in *APOE* genotype groups

When the whole brain image used as input, MAE ranged from 2.62 to 2.94 years (Table 2). *APOE* ε4 homozygotes (ε4/ε4) showed greater whole brain age gap (β = 0.162, CI = [0.081, 0.244], p = 9.98e-05). Details can be found in Table S2.

#### Associations between HA gap and cognition

In the cross-sectional analysis in UK Biobank, 19,441 participants have both HA gap data and cognition data. The greater HA gap in both left and right hemispheres was associated with lower cognitive performance: processing speed, executive function, memory, and global cognition. Detailed results can be found in Figure 3.

**Figure 3.**
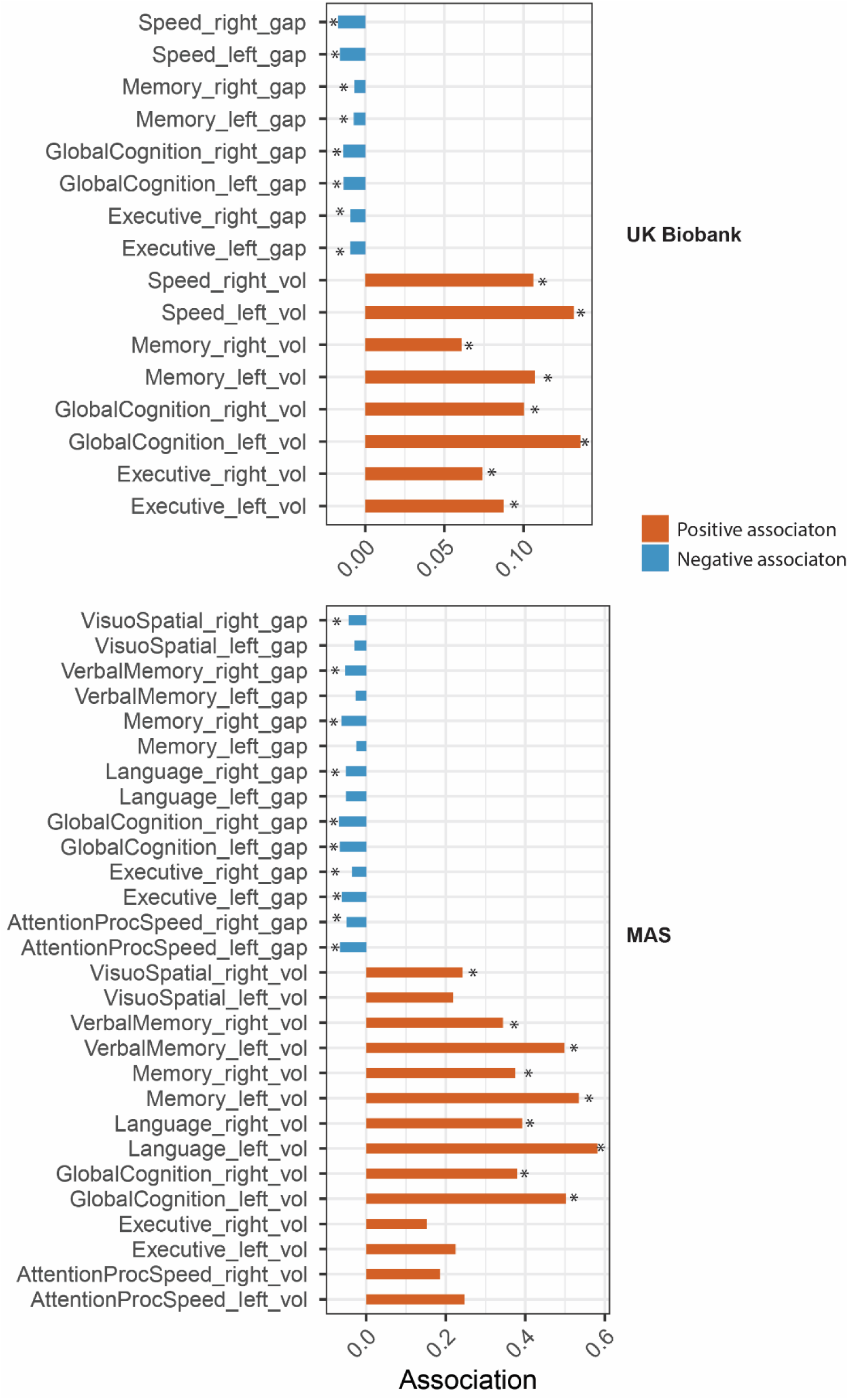
Associations between HA gap (or hippocampus volume) and cognition. Here, “gap” refers to the HA gap, while “vol” denotes hippocampus volume. Asterisks indicate statistically significant associations after false discovery rate (FDR) correction (p<0.05). MAS: Sydney Memory and Ageing Study.

#### HA gap and modifiable risk factors

As both the left and right HA gaps were significantly associated with non-modifiable variable: *APOE* genotype, we continued to examine their associations with modifiable risk factors: (hypertension, diabetes, alcohol intake frequency, smoking status, and physical activity). In UK Biobank dataset, we found that participants diagnosed with hypertension or diabetes showed significantly greater HA gap compared with healthy counterparts. Additionally, participants who have alcohol 1-3 times a month or special occasion only showed a smaller HA gap compared with daily drinkers (p<0.001). Previous and current smokers exhibited a significantly greater HA gap compared with non-smokers (p<0.001). However, we did not find significant associations between HA gap and physical activities. All details can be found in Figure 4 and Table S3.

**Figure 4.**
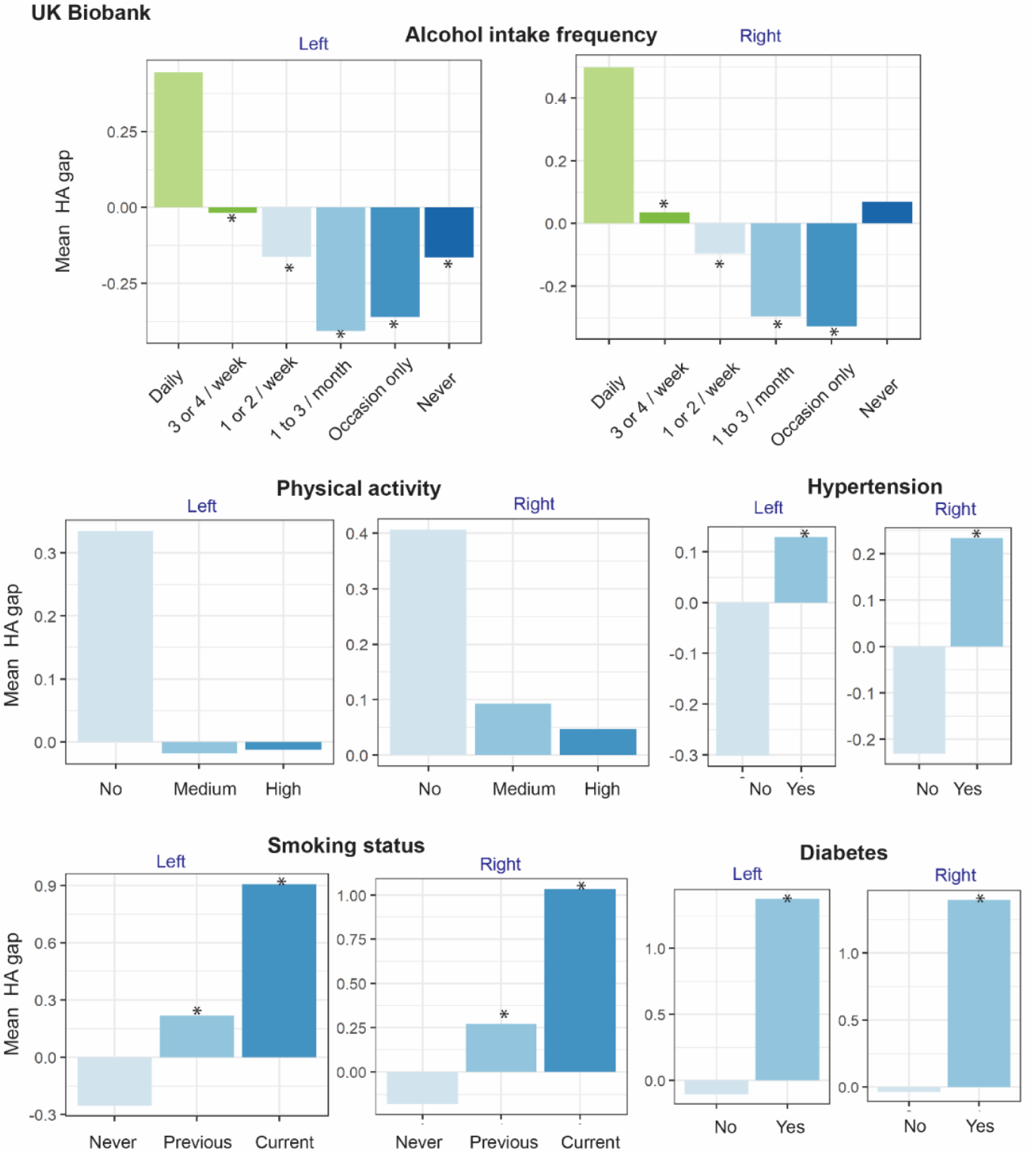
Associations between mean HA gap and other variables in UK Biobank. The X axis represents different groups based on different variables. The Y axis represents the mean left and right HA gap within each group. In each variable category, the first group served as the reference, and subsequent groups were compared with their corresponding reference group. Groups displaying statistically significant differences compared to the reference group were marked with an asterisk (*). Details can be found in Table S3. HA: hippocampus ROI age.

### 3.3 Longitudinal analysis

#### The HA gap change rate in *APOE* genotype groups

Compared with *APOE* ε2 homozygotes, *APOE* ε4 homozygotes showed a greater longitudinal change rate in the left HA gap (β = 0.581, p = 0.025) (Table 4 and Figure 2). However, this effect was not observed in the right hemisphere. The left HA gap trajectory in different *APOE* genotype groups can be found in Figure S2. In addition, no significant differences in annual HV changes were found among the various *APOE* genotype groups.

**Table 4:**
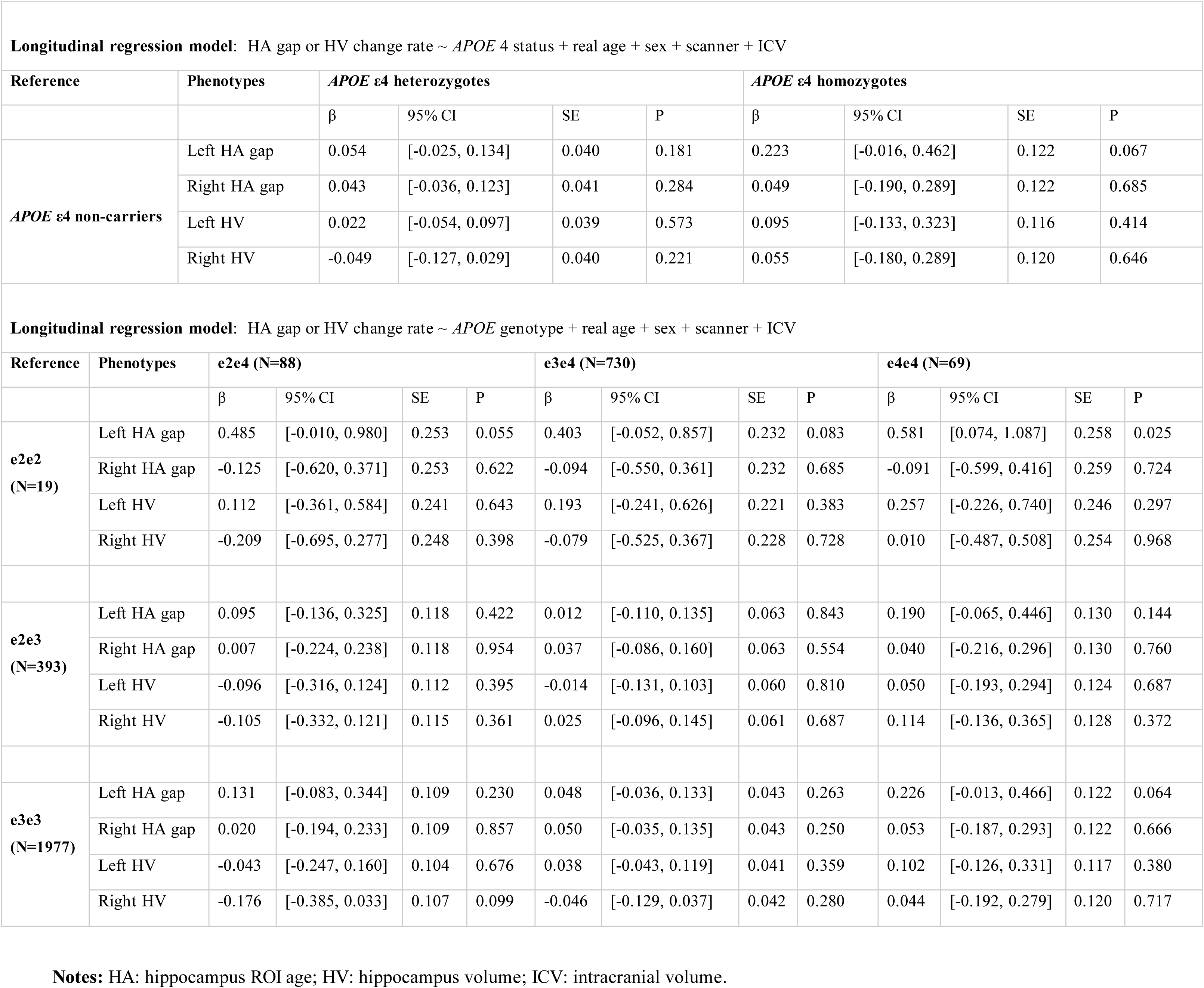
Longitudinal analysis in UK Biobank: The change rate of HA gap and HV in different *APOE* genotype groups.

### 3.4 Occlusion analysis

As a greater longitudinal age gap change rate in *APOE* ε4 homozygotes was only found in the left hippocampus ROI (compared with *APOE* ε2 homozygotes), we subsequently performed occlusion analysis on this region. The saliency map of the prediction model revealed that beyond the hippocampus, regions in proximity, including peaks around the thalamus, pallidum, putamen, amygdala, nearby cerebral cortex, and cerebral white matter, also play an important role in age prediction (Figure 5). The detailed saliency map can be found in Figure S3. Additionally, compared with the original model performance, the model using ROI without hippocampus exhibited higher MAE (2.96 – 3.59 years in left HA model; 2.79 – 3.50 years in right HA model). The specific MAE values for each of the five folds can be found in Table 2.

**Figure 5.**
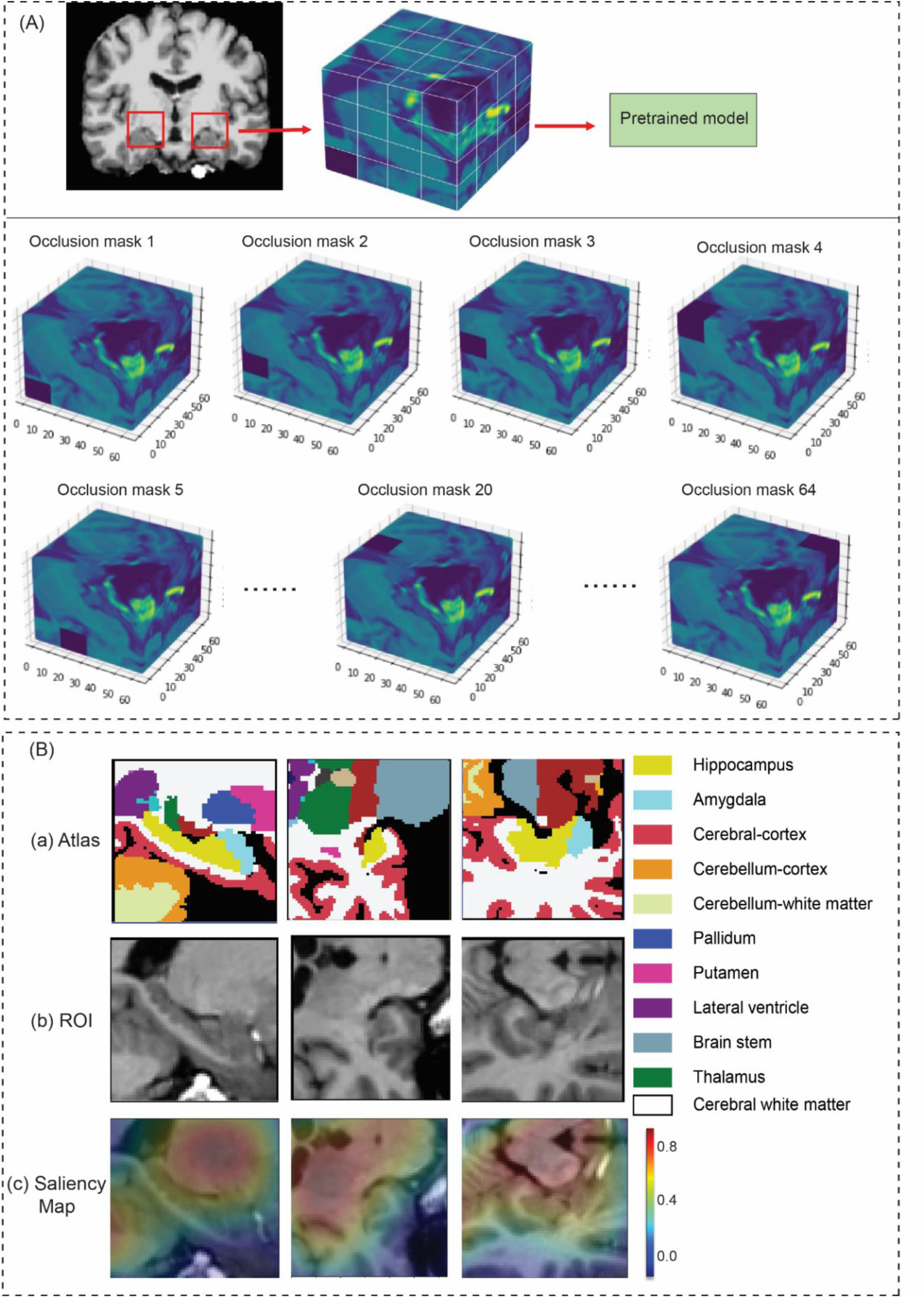
The occlusion analysis. (A) The occlusion analysis framework and the illustration of the different occlusion mask. (B) visualization of hippocampus ROI. The saliency map was produced by occlusion analysis. The warmer colours represent the higher importance of a region in HA prediction. HA: hippocampus ROI age.

### 3.5 Transfer learning result in MAS dataset

The pretrained models from the UK Biobank were applied to the MAS dataset using transfer learning, and the performance is summarized in Table 5. The models were evaluated using three-fold cross-validation in MAS, with the MAE calculated for each test fold. For the left hippocampus ROI, the MAE ranged from 3.12 to 3.86 years. For the right hippocampus ROI, the MAE ranged from 3.11 to 4.77 years. These results indicated that the pretrained models from the UK Biobank can be effectively transferred to the MAS dataset, though the performance is generally lower than in the original UK Biobank dataset. The associations between HA gap, *APOE* genotypes, cognition, and modifiable risk factors can be found in Figure 2, Figure 3, and Figure S4.

**Table 5.** Performance of pretrained model applied to MAS using transfer learning.

| MAS measures | Pre-trained model | MAS: CV1 | MAS: CV2 | MAS: CV3 |
| --- | --- | --- | --- | --- |
| **Left ROI (MAE)** | UKB-CV1 | 3.31 | 3.13 | 3.47 |
|  | UKB-CV2 | 3.45 | 3.15 | 3.86 |
|  | UKB-CV3 | 3.37 | 3.12 | 3.56 |
|  | UKB-CV4 | 3.46 | 3.20 | 3.48 |
|  | UKB-CV5 | 3.38 | 3.14 | 3.62 |
| **Right ROI (MAE)** | UKB-CV1 | 3.50 | 3.23 | 4.30 |
|  | UKB-CV2 | 3.64 | 3.11 | 4.55 |
|  | UKB-CV3 | 3.56 | 3.14 | 4.59 |
|  | UKB-CV4 | 3.53 | 3.13 | 4.77 |
|  | UKB-CV5 | 3.50 | 3.11 | 4.41 |

**Notes:** MAE: mean absolute error. CV: cross-validation. UKB-CV1-CV5: five-fold cross-validation in UK Biobank. MAS:CV1-CV3: three-fold cross-validation in MAS. ROI: regions of interest. MAS: Sydney Memory and Ageing Study.

## 4 Discussion

In the current study, we developed a 3D-CNN model to predict the age of left and right hippocampus ROI in a large population sample. Subsequently, we evaluated the *APOE* genotype influences on the HA gap, the change rate of HA gap, cognition, and various modifiable risk factors. Our findings indicate that *APOE* ε4 influences the HA gap in both hemispheres, and individuals with greater HA gap tend to exhibit poorer cognitive performance. Specially, participants with hypertension, diabetes, those who engage in heavy daily alcohol consumption, or are previous/current smokers are more likely to have a greater HA gap, which serves as an indicator of poorer health. In the longitudinal analysis, a significant difference in the change rate of the left HA gap was observed between *APOE* ε4 homozygotes and *APOE* ε2 homozygotes.

In the cross-sectional analysis, the HA gap varied across different *APOE* genotype groups, with *APOE* ε4 homozygotes (*APOE* ε4/ε4 carriers) exhibiting the greatest average HA gap in both hemispheres compared to others (ε2/ε2, ε2/ε3, ε3/ε3). This suggests that *APOE* ε4/ε4 accelerates the ageing process in the hippocampus ROI. In contrast, *APOE* ε4 heterozygotes showed a significantly greater HA gap only in right hemisphere compared to *APOE* ε4 non-carriers. Additionally, the differences in hippocampal volume (HV) across *APOE* genotypes were less pronounced than the differences in HA gap, indicating that *APOE* genotype may have a stronger influence on HA gap than on HV. Our findings indicated that *APOE* ε4 carriers exhibit not only smaller hippocampus volume but also a greater HA gap, even in the absence of an AD diagnosis.

Advanced age is usually accompanied by cognitive decline, and studies have consistently indicated a negative association between brain age gap and cognitive scores (Cole et al., 2018; Elliott et al., 2021). The brain age gap may act as the mediator between modifiable risk factors and cognitive functioning (Chen et al., 2022). In early onset Alzheimer’s disease, *APOE* ε4 carriers tend to experience a more rapid decline in cognitive functions such as processing speed, executive function, and memory (Polsinelli et al., 2023). Our finding that a greater HA gap is associated with poorer cognitive performance adds to the body of evidence supporting the link between brain age gap and cognitive decline.

Interestingly, in the longitudinal analysis, a notable divergence was observed: *APOE* ε4/ε4 carriers showed a rapid increase in left HA gap, while ε2/ε2 carriers demonstrated a decreased HA gap over the years. These effects, however, were not mirrored in the change rate of the right HA gap. Given the established higher risk associated with *APOE* ε4/ε4 for AD and the protective nature of *APOE* ε2/ε2 (Serrano-Pozo et al., 2021), it is plausible that the increased change rate of HA gap contributes to the heightened risk in ε4/ε4 carriers, particularly in the left hippocampus ROI. Supporting this, a recent study demonstrated that *APOE* ε4 contributed to faster-accelerated atrophy in the left hippocampus in persistent cognitive normal and the transition from normal cognitive stages to dementia (Huang et al., 2023). Furthermore, verbal memory decline was also correlated with left hippocampal atrophy (Baxter et al., 2023). In addition to longitudinal volume atrophy, *APOE* ε4 was also related to longitudinal decrease in hippocampal activation in normal ageing (Håglin et al., 2023). Our observation of longitudinal change rate in HA gap provides additional evidence that *APOE* ε4 can influence both functional and structural aspects of hippocampus-related regions.

The investigation into associations between HA gaps and both non-modifiable and modifiable variables has yielded insightful findings. Participants diagnosed with hypertension or diabetes exhibited significantly greater HA gaps compared to their healthier counterparts. Additionally, individuals who engage in heavy daily alcohol consumption or smoking also exhibited greater HA gaps. This aligns with previous findings that link increased brain age to frequent tobacco and alcohol use (Ning et al., 2020). The association between unhealthy lifestyle, such as high alcohol use, and lower hippocampal volume (Binnewies et al., 2023) may further explain why heavy daily alcohol users tend to have a larger HA gap. Together, these findings enhance our understanding of the complex factors contributing to aging in hippocampus-related regions.

The “black box” nature of deep learning models usually makes the results hard to explain, but occlusion analysis in the current study helped the interpretability of the HA prediction model. Interestingly, occlusion analysis demonstrated that the hippocampus nearby regions have great contribution to HA prediction. Specifically, thalamus, pallidum, nearby cerebral cortex, and cerebral white matter are critical regions in HA prediction. Many of these regions belong to the basal ganglia and cerebellum, which are essential for motor control and emotional processing and influence limbic function (Arber and Costa, 2022; Pierce and Péron, 2020). Age-related differences in thalamocortical connectivity can contribute to changes in attention, working memory, and episodic memory processes associated with ageing (Fama and Sullivan, 2015; Hughes et al., 2012; Ystad et al., 2010). Additionally, the HA prediction model using 64×64×64 ROI removing hippocampus showed a slightly larger MAE than the original model which included hippocampus. This slight MAE differences between models indicate that while the hippocampus contributes to HA prediction, the surrounding regions also play a vital role in age estimation. Collectively, these analyses demonstrate that the 3D bounding boxes of the ROI surrounding the hippocampus could serve as a valuable representation for brain age prediction.

Our study has some strengths and limitations. Firstly, while the sample size for both training and test sets is relatively large, generalizability to broader populations, including diverse demographics and clinical conditions, may require further exploration. Secondly, although our findings offer valuable insights into the associations between HA gap and factors such as *APOE* genotype, chronic diseases, and lifestyle factors, it is important to note that our study does not establish HA gap as a definitive biomarker for AD. Future research is needed to thoroughly investigate and validate the potential of HA gap as a biomarker for AD diagnosis, particularly in cohorts with confirmed AD diagnoses. Lastly, while our study focused on associations with *APOE* genotype, chronic diseases, and lifestyle factors, incorporating additional variables such as other genetic markers, socioeconomic factors, and broader health metrics could offer a more comprehensive view of the determinants influencing HA gap.

## Conclusion

In conclusion, our study has shed light on the intricate relationship between HA gap and various determinants, including both non-modifiable factors like *APOE* genotype and modifiable factors. *APOE* ε4 homozygotes exhibited greater left and right HA gaps, along with a faster change rate in left HA gap, indicating that left HA gap might be a potential *APOE*-related biomarker or a health indicator.

## Data Availability

All data produced in the present study are available upon reasonable request to the authors.

## Acknowledgments

C.D. is supported by Scientia Scholarship of University of New South Wales, Sydney. We extend our gratitude to the UK Biobank and the Sydney Memory and Ageing Study (MAS) databases for providing valuable data.

