## Supplemental Files for "Deep learning-derived age of hippocampus-centred regions is influenced by *APOE* genotype and modifiable risk factors"

**Supplementary Materials**

| Disease | Code |
| --- | --- |
| Stroke or ischaemic stroke | 1081 |
| Transient ischaemic attack | 1082 |
| Subdural haematoma | 1083 |
| Subarachnoid haemorrhage | 1086 |
| Neurological injury/trauma | 1240 |
| psychological/psychiatric problem | 1243 |
| Infections of the nervous system | 1244 |
| Brain/intracranial abscess | 1245 |
| Encephalitis | 1246 |
| Meningitis | 1247 |
| Guillain-Barré syndrome | 1256 |
| Chronic degenerative neurological | 1258 |
| Motor Neuron Disease | 1259 |
| Multiple Sclerosis | 1261 |
| Parkinson’s disease | 1262 |
| Dementia or Alzheimer’s disease | 1263 |
| Epilepsy | 1264 |
| Head injury | 1266 |
| depression | 1286 |
| schizophrenia | 1289 |
| mania/bipolar disorder/manic depression | 1291 |
| Other demyelinating disease | 1397 |
| Cerebral aneurysm | 1425 |
| Cerebral palsy | 1433 |
| Brain haemorrhage | 1491 |
| Spina bifida | 1524 |
| Ischaemic stroke | 1583 |
| Meningioma (benign) | 1659 |

### Table S1: UK Biobank exclusive disease and corresponding codes

**Notes**: participants in UK Biobank with any diseases listed above were remove.

### Table S2: The whole brain age gap in different APOE genotype groups

The original whole brain image had dimensions of 166 × 166 × 189 voxels with a resolution of 1 × 1 × 1 mm. To optimize the model's computational efficiency, we resampled the image to 83 × 83 × 94 voxels with a resolution of 2 × 2 × 2 mm. We employed a similar 3D-CNN architecture with a learning rate of 1×10⁻³ and a batch size of 56. The associations between the whole brain age gap and *APOE* genotype are presented in Table S2.

| **Cross-sectional regression model**: Whole brain age gap ~ *APOE* 4 status + real age + sex + scanner + ICV | | | | | | | | | | | | | | | | | | | | | | |
| --- | --- | --- | --- | --- | --- | --- | --- | --- | --- | --- | --- | --- | --- | --- | --- | --- | --- | --- | --- | --- | --- | --- |
| **Reference** | |  | | | ***APOE*** **ε4** **heterozygotes** | | | | | | | | | ***APOE*** **ε4** **homozygotes** | | | | | | | | |
| ***APOE*** **ε4 non-carriers** | | Whole brain | | | β | 95% CI | | | | SE | | P | | β | | 95% CI | | | SE | | P | |
|  |  |  |  |  | 0.021 | [-0.006,0.049] | | | | 0.014 | | 0.130 | | 0.162 | | [0.081,0.244] | | | 0.042 | | 9.98e-05 | |
| **Cross-sectional regression model**: Whole brain age gap ~ *APOE* genotype + real age + sex + scanner + ICV | | | | | | | | | | | | | | | | | | | | | | |
| **Reference** | **Phenotypes** | | **e2e4** | | | | | | **e3e4** | | | | | | | | **e4e4** | | | | | |
|  |  | | β | 95% CI | | | SE | P | β | | 95% CI | | SE | | P | | β | 95% CI | | SE | | P |
| **e2e2** | Whole brain | | -0.051 | [-0.237,0.123] | | | 0.095 | 0.588 | 0.018 | | [-0.152,0.187] | | 0.086 | | 0.839 | | 0.152 | [-0.034,0.338] | | 0.095 | | 0.108 |
| **e2e3** |  |  | -0.042 | [-0.129,0.044] | | | 0.044 | 0.337 | 0.027 | | [-0.016,0.069] | | 0.022 | | 0.219 | | 0.161 | [0.074,0.249] | | 0.045 | | 3.02e-04 |
| **e3e3** |  |  | -0.041 | [-0.122,0.040] | | | 0.041 | 0.320 | 0.028 | | [-0.002,0.057] | | 0.015 | | 0.063 | | 0.163 | [0.081,0.245] | | 0.042 | | 1.02e-04 |

Table S2: The whole brain age gap in different APOE genotype groups

**Notes:** ICV: intracranial volume.

### Table S3: Associations between HA gap and modifiable risk factors in UK Biobank.

| **Reference** | **Risk factors** | **β** | | **SE** | | **P** | |
| --- | --- | --- | --- | --- | --- | --- | --- |
|  |  | Left HA gap | Right HA gap | Left HA gap | Right HA gap | Left HA gap | Right HA gap |
| Non-hypertension | Hypertension | 0.0840 | 0.0953 | 0.0155 | 0.0155 | 6.29e-08 | 8.63e-10 |
| Non-diabetes | Diabetes | 0.3223 | 0.3392 | 0.0350 | 0.0351 | 3.94e-20 | 4.38e-22 |
| Alcohol intake daily or almost daily | Alco_2 | -0.0885 | -0.1017 | 0.0206 | 0.0206 | 1.75e-05 | 8.25e-07 |
|  | Alco_3 | -0.1315 | -0.1410 | 0.0215 | 0.0215 | 9.74e-10 | 5.85e-11 |
|  | Alco_4 | -0.2090 | -0.2143 | 0.0278 | 0.0279 | 6.10e-14 | 1.51e-14 |
|  | Alco_5 | -0.1917 | -0.2035 | 0.0320 | 0.0320 | 2.18e-09 | 2.19e-10 |
|  | Alco_6 | -0.1250 | -0.0472 | 0.0390 | 0.0390 | 0.0014 | 0.2269 |
| Never smoke | Previous smoker | 0.0864 | 0.0873 | 0.0162 | 0.0162 | 9.48e-08 | 7.25e-08 |
|  | Current smoker | 0.2786 | 0.3010 | 0.0322 | 0.0323 | 6.05e-18 | 1.20e-20 |
| No physical activity | Medium level physical activity | -0.1349 | -0.0525 | 0.0692 | 0.0693 | 0.0513 | 0.4484 |
|  | High level physical activity | -0.1233 | -0.0669 | 0.0662 | 0.0662 | 0.0625 | 0.3126 |

**Notes**: regression model: left or right HA gap ~ hypertension + diabetes + alcohol intake frequency + smoking status + physical activity + BMI + *APOE* ε4 status + baseline age + sex + scanner + ICV. HA: hippocampus ROI age. Alco_2: alcohol intake three or four times a week. Alco_3: once or twice a week. Alco_4: one to three times a month. Alco_5: special occasion only. Alco_6: never.

### Figure S1: CNN performance on the five-fold test dataset in UK Biobank

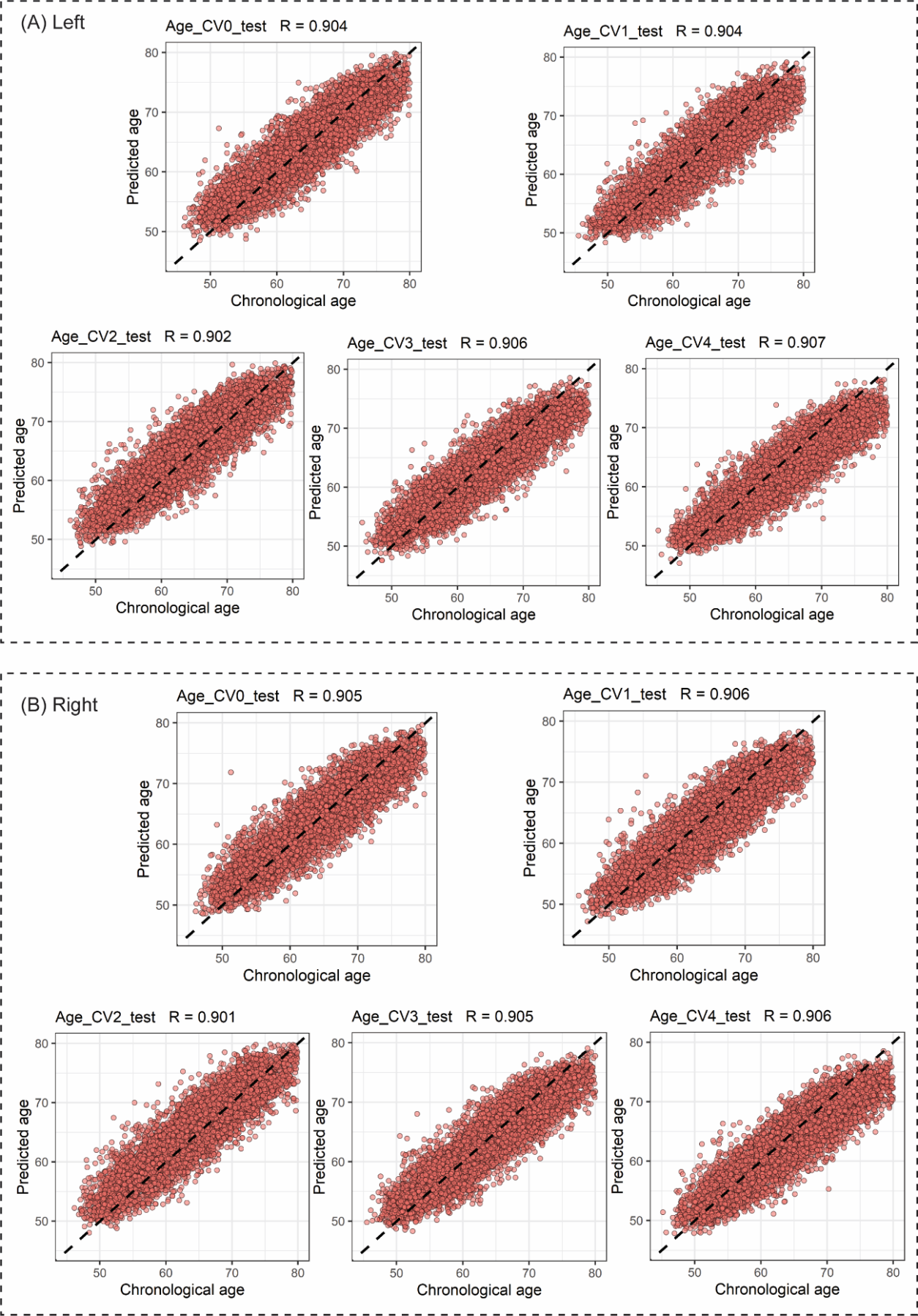

**Figure S1. CNN performance on the five-fold test dataset in UK Biobank.** (A) The plots depict chronological age (x-axis) and left HA age (y-axis) with Pearson correlation. (B) The plots depict chronological age (x-axis) and right HA age (y-axis) with Pearson correlation.

### Figure S2: The left HA gap trajectory by chronological age in different *APOE* genotype groups

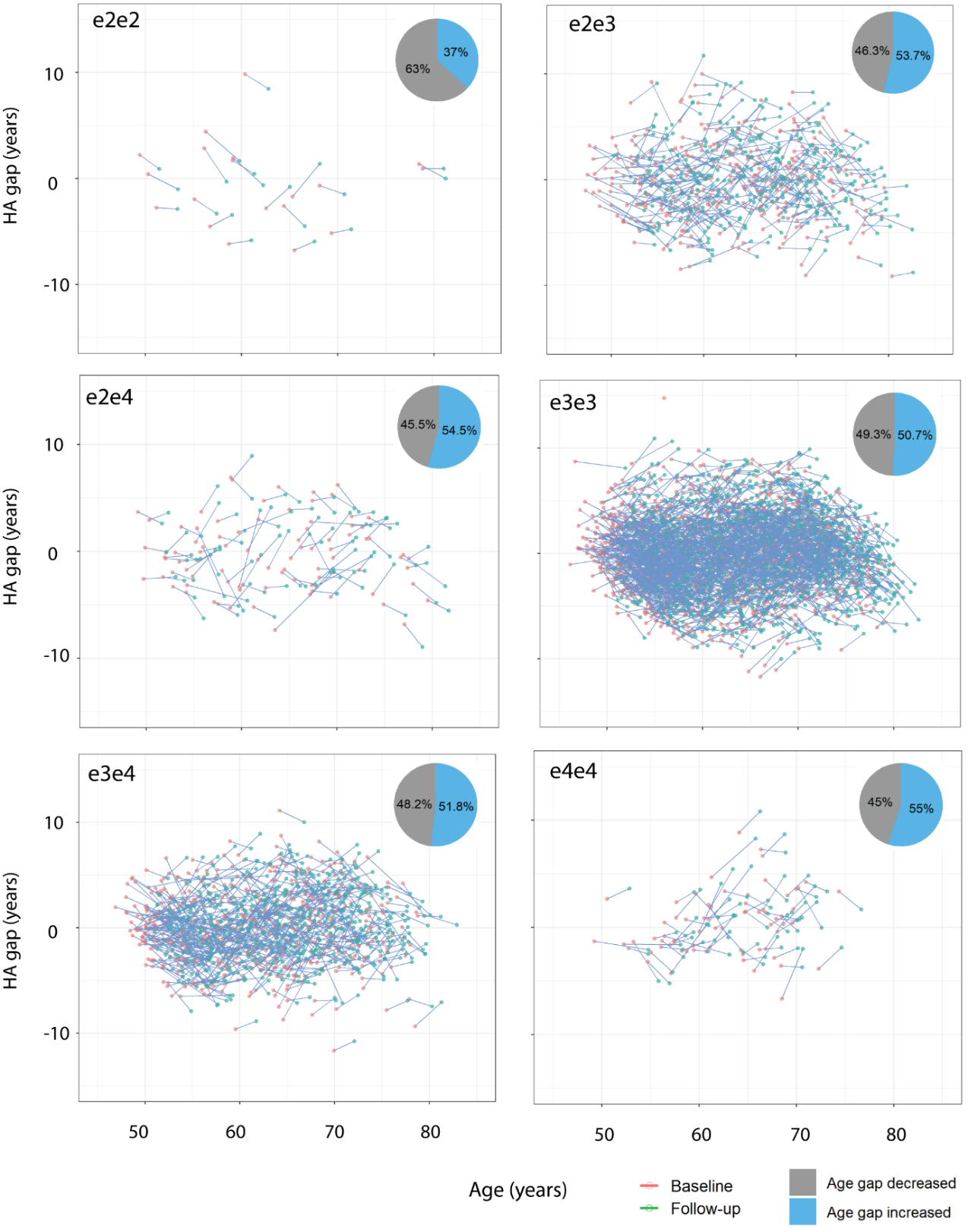

**Figure S2. The left HA gap trajectory by chronological age in different *APOE* genotype groups.** Connected points represent baseline HA gap and follow-up HA gap for an individual. The pie plot in the upper right corner represents the percentage of participants who have decreased HA gap (grey) and increased HA gap (blue) during follow-up.

### Figure S3: The ROI atlas and saliency map

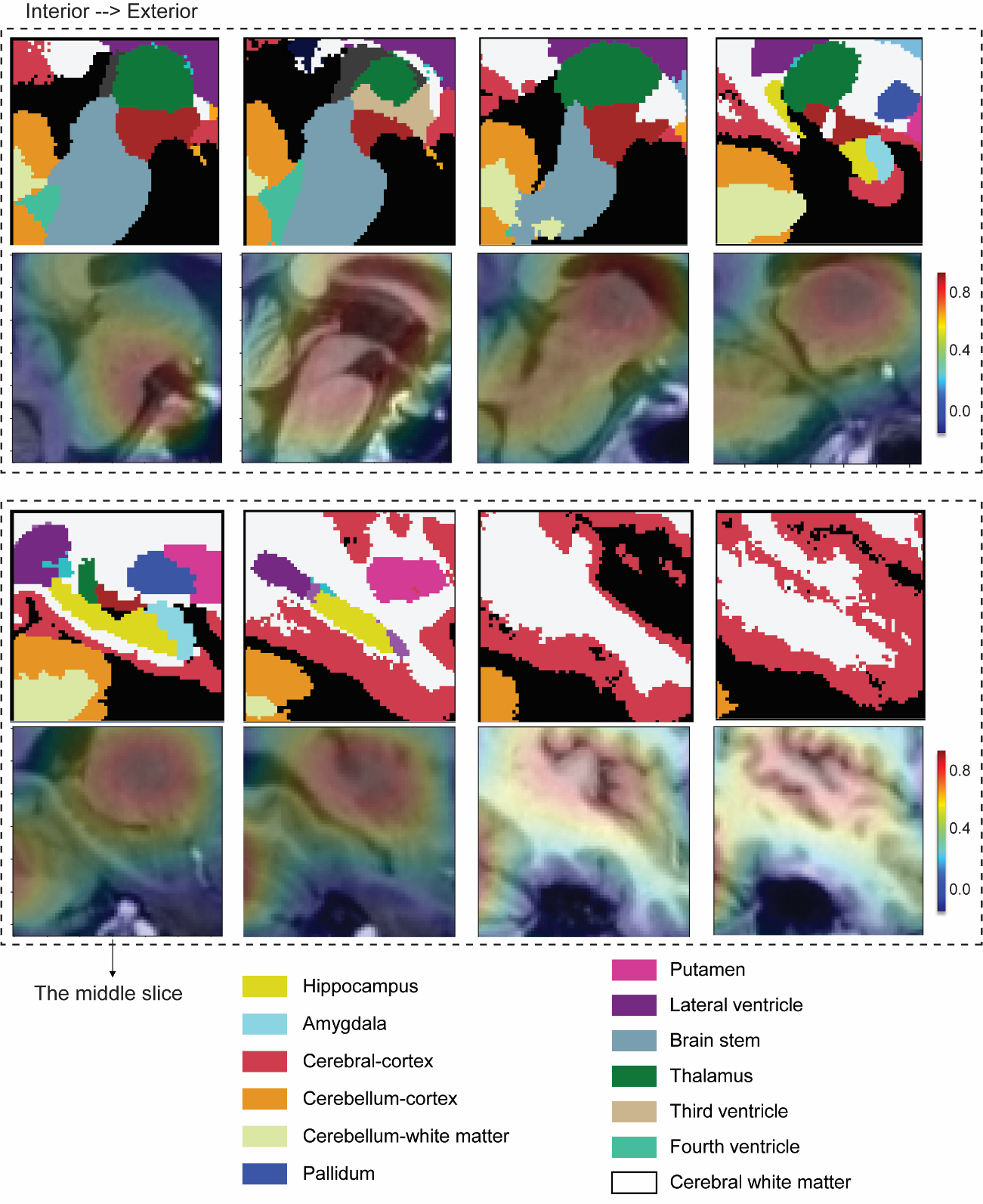

**Figure S3. The ROI atlas and saliency map**. The warmer colours represent the higher importance of a region in HA prediction.

### Figure S4: Associations between mean HA gap and other variables in MAS

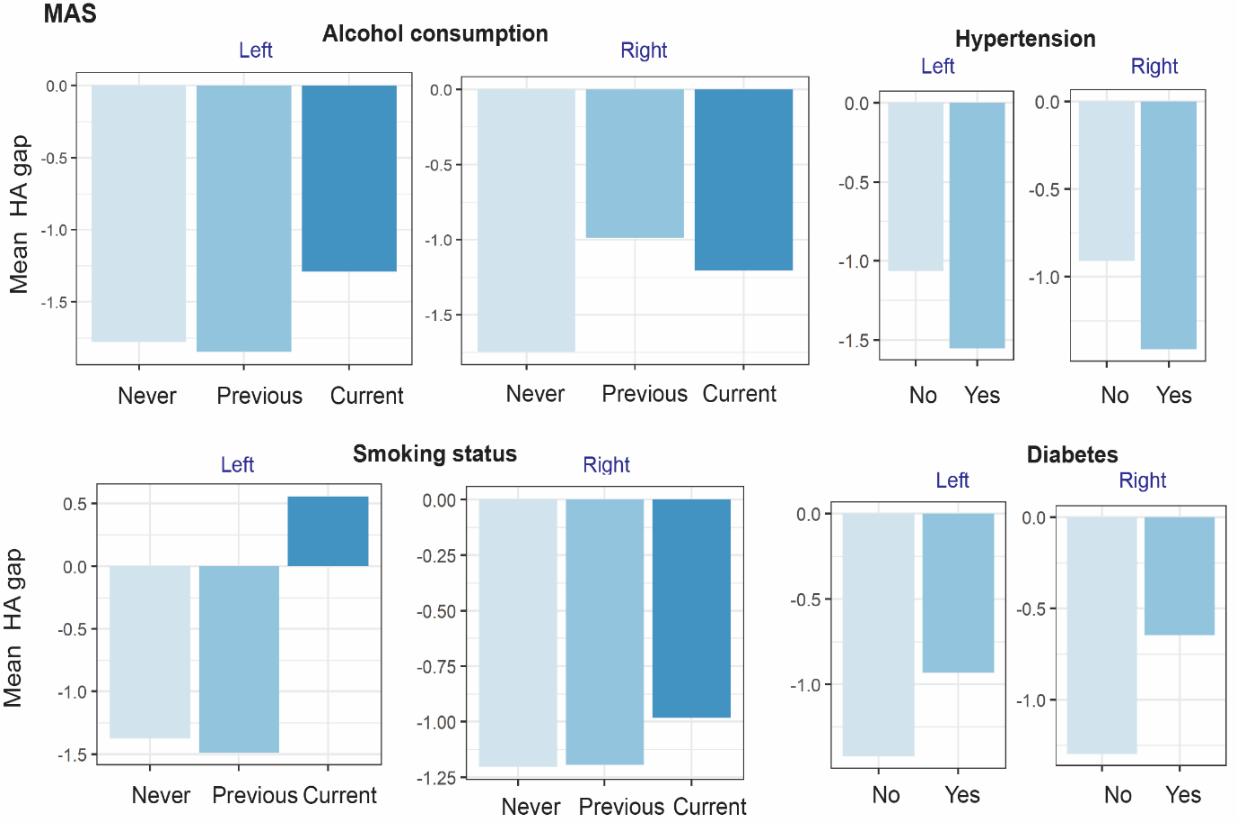

**Figure S4. Associations between mean HA gap and other variables in MAS**. The X axis represents different groups based on different variables. The Y axis represents the mean left and right HA gap within each group. In each variable category, the first group served as the reference, and subsequent groups were compared with their corresponding reference group. Groups displaying statistically significant differences compared to the reference group were marked with an asterisk (*).
